# Water, Sanitation, Hygiene (WASH) or others? - a global analysis on determinants of Covid-19 pandemic

**DOI:** 10.1101/2020.08.11.20173179

**Authors:** Ajishnu Roy, Aman Basu, Kousik Pramanick

## Abstract

Coronavirus (SARS Covid-19) has become a global public health concern due to its unpredictable nature and lack of adequate pre-existing conditions. Achievement of WASH services is being acknowledged as indispensable in safeguarding health. However, on a global scale, it is currently not clear whether deprivation or non-obtainability of which of the factors are closely related to Covid-19 dynamics and up to which degree. We have analysed 6 months’ data related to five Covid-19 indicators for most of the countries in the world with three groups of indicators of WASH, socio-economic factors and mobility & stringency. Four successive steps were followed to carry out the analysis: (a) identification of relevant key explanatory variables of Covid-19, (b) identification of indicators (n=103) for other 5 sub-groups of factors (viz. WASH, economy, health, society, mobility & stringency) through literature review, (c) utilizing Spearman’s rank-order correlation for measuring the association of the possible explanatory variables with those of Covid-19 (of spatial nature), and then (d) continuing the same for 6 consecutive months (from March to August 2020) until we get an almost unchanging trend. We have found a strong positive correlation between lesser effects (i.e. either less confirmed case or higher recovery rate) of Covid-19 and better access to WASH as well as socioeconomic factors throughout this time for a significant amount of the indicators. Since a handful of research is available to discuss the association between Covid-19 with other probable determinants along with the nature and degree of their relationship, this study should be perceived as an expanded view on the complexities of Covid-19 interrelationships towards understanding determinants of Covid-19 dynamics, which could help to shape an agenda for research into some unanswered questions.

## 1. Introduction

Severe acute respiratory syndrome-coronavirus (SARS-CoV-2), emerged in China in December 2019, is causing an outbreak of respiratory disease (COVID-19 disease by WHO). The new SARS-CoV-2 virus is known to spread by person-to-person contact (through respiratory droplets over a short distance) or via faecal-oral routes (hypothesised by Heller et al. 2020). Studies have shown that coronaviruses exist and can maintain their viability in sewage and hospital wastewater, originating from the faecal discharge of infected patients. Concerns have been raised over inequity in access to various prevention and control measures for slum dwellers, refugees etc. (Singh et al. 2020), even though Tedros Adhanom Ghebreyesus (DG, WHO) has urged a “whole-of-government, whole-of-society approach” for Covid-19 (Lau et al. 2020). According to Lau et al. (2020), one of the important reasons for this might be limited access to safe water and sanitation. Connection of control of covid-19 and global access to handwashing facilities has also been established for low-income countries (Brauer et al. 2020). The insecurity of water may also act as a deterrent for covid-19 mitigation, especially in developing areas (Stoler et al., 2020). Odith et al. (2020) have also emphasized probable induction of Covid-19 transmission rate from the shortfall in water and sanitation, using Nigeria as a case study. Amankwaa & Fischer (2020) and Jiwani & Antiporta (2020) have also emphasised on the effects of poor WASH services on Covid-19 fatalities in sub-Saharan African countries. Caruso & Freeman (2020) have argued on the effects of inequality of sanitation facility with relation to the spread of Covid-19. Mushi & Shao (2020) have stated the possible role of WASH services regarding the prevention of the further spread of covid-19, especially for lower-and middle-income countries. Ray (2020) have opined on the importance of hygiene about Covid-19. All of these works indicate a possible significant level of connection between inequality of WASH services and Covid-19. WHO and UNICEF have long been suggested about the importance of WASH in itself (UNICEF 2019) and also related to Covid-19 (WHO, 2020 a, b). Thus, these works indicate towards shortfalls WASH may relate to higher Covid-19 (occurrence and transmission).

Some socio-economic and demographic parameters were closely associated with COVID-19 cases-fatality rate. These include - per capita gross national income, net migration, voters’ turnout ratio for Asian countries (Varkey et al. 2020), average age, the population density and density of doctors in Germany (Ehlert, 2020), tourism (Farzanegan et al. 2020; Lapointe, 2020; Gössling et al. 2020), household size, use of public transport, aged population and self-reported health for England and Wales (Sá, 2020), mean income in the 10 districts of the city of Barcelona – Spain (Baena-Díez et al., 2020), socioeconomic characteristics in neighbourhood community in 7 states of USA (Hatef et al., 2020), early life environment, life trajectory and socioeconomic status (Holuka et al. 2020), income and population density (Jani & Mavalankar, 2020), morbidity rate, risky health behaviours, crowding, and population mobility in South Korea (Weinstein et al. 2020), poor healthcare infrastructure, high population and room density, and informal job markets in low-income countries (UN Habitat, 2020; Mishra et al., 2020), poverty and income inequality in ten Latin America and the Caribbean (LAC) cities (Bolaño-Ortiz et al., 2020), working age population (i.e. demographic composition) in Italy and South Korea (Dowd et al., 2020), total population, poverty and income for some of the 31 countries in Europe (Sannigrahi et al., 2020), population, GDP and population density for Wuhan, China (Xiong et al., 2020) etc. These studies indicate a varied relationship between social, economic, demographic influencing factors and Covid-19.

Apart from these, there are, at least 2 groups of real-time indicators which are supposed to have intervened with the dynamic of Covid-19, one is government measures through lockdowns and containment protocols and another is people’s daily behaviour and activities. First, has mostly been interpreted through Oxford COVID-19 Government Response Tracker (OxCGRT) (Hale et al. 2020) and the second through Google community mobility reports (Google, 2020). Some works have suggested in support of their association with Covid-19 (Carlitz and Makhura, 2020; Castex et al., 2020; Yang et al., 2020).

There are some drawbacks to these works. Either some studies have incorporated a selectively few arrays of indicators, or their operational boundary is one or a group of countries of a region. Also, most of them deal with Covid-19 dataset of one single date, whereas it is well known that various parameters of this pandemic are very spatiotemporally dynamic i.e., changing with time as well as space (Sahin et al., 2020). For these reasons, we have conceptualized this work.

## 2. Research Questions

There are some features that we have tried to encapsulate in this study.

(a) Are the various indicators of Covid-19 and indicators of WASH-related, at the global scale?
(b) If they are related, how much is their degree of association?
(c) Does their relationship remain similar throughout Covid-19 pandemic? i.e. Do the WASH conditions are affecting covid-19 conditions similarly from the onset of the pandemic?
(d) If the effect of WASH on indicators of Covid-19 have decreased over time, then what about other socioeconomic factors?
(e) Are the various indicators of socioeconomic dimensions and indicators of WASH-related, at the global scale?
(f) If socioeconomic factors and Covid-19 are related, how much is their degree of association?
(g) Do the socioeconomic factors are affecting covid-19 conditions similarly from the onset of the pandemic?
(h) If the effects of socioeconomic factors (excluding WASH) on indicators of Covid-19 have decreased over time, then what about stringency and mobility factors?
(i) If the stringency and mobility factors are related to Covid-19, how much is their degree of association?
(j) Do the stringency and mobility factors affect covid-19 conditions similarly from the onset of the pandemic?

## 3. Method and data

We have collected data of water, sanitation and hygiene from WHO/UNICEF JMP (2017) dataset and WDI (2020). We have used the data from the latest available year in this work. These set of indicators have been used as indicators of socioeconomic status following the framework used by Roy and Pramanick (2019). We have collected data of Covid-19 from for 6 different dates (30^th^ of March to 30^th^ August, 2020) from JHU CCSE (2020). We have used data for 5 dimensions of water, 5 dimensions of sanitation, 3 dimensions of hygiene (i.e. 63 indicators of WASH, suppl. table 1-3) and 5 dimensions (with 5 indicators) of Covid-19. Also, we have used data for 21 dimensions (via 33 indicators) of socio-economy (except – WASH, suppl. table 4) and 2 dimensions (with 7 indicators, suppl. table 5) of stringency and mobility. List of indicators is given in table 1. More details about sub-indicators are provided in the supplementary file.

**Table 1:**
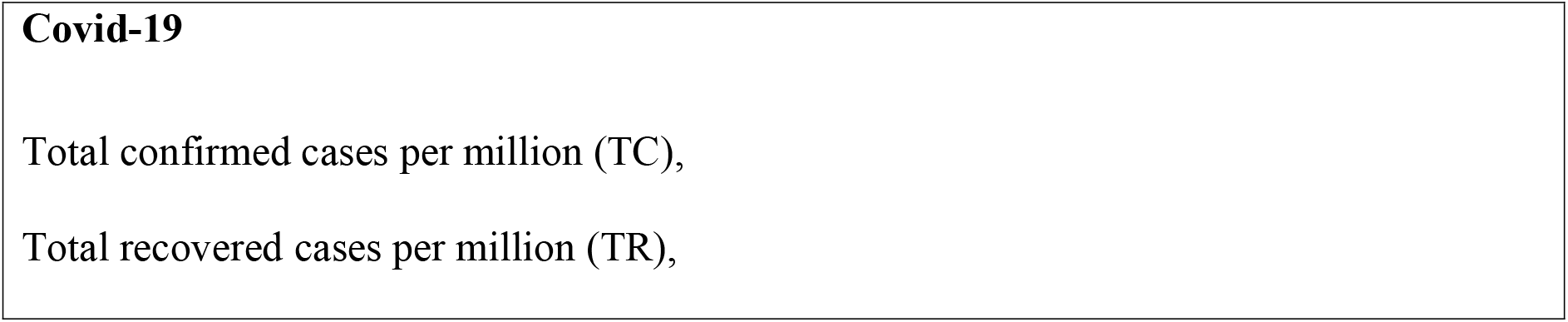
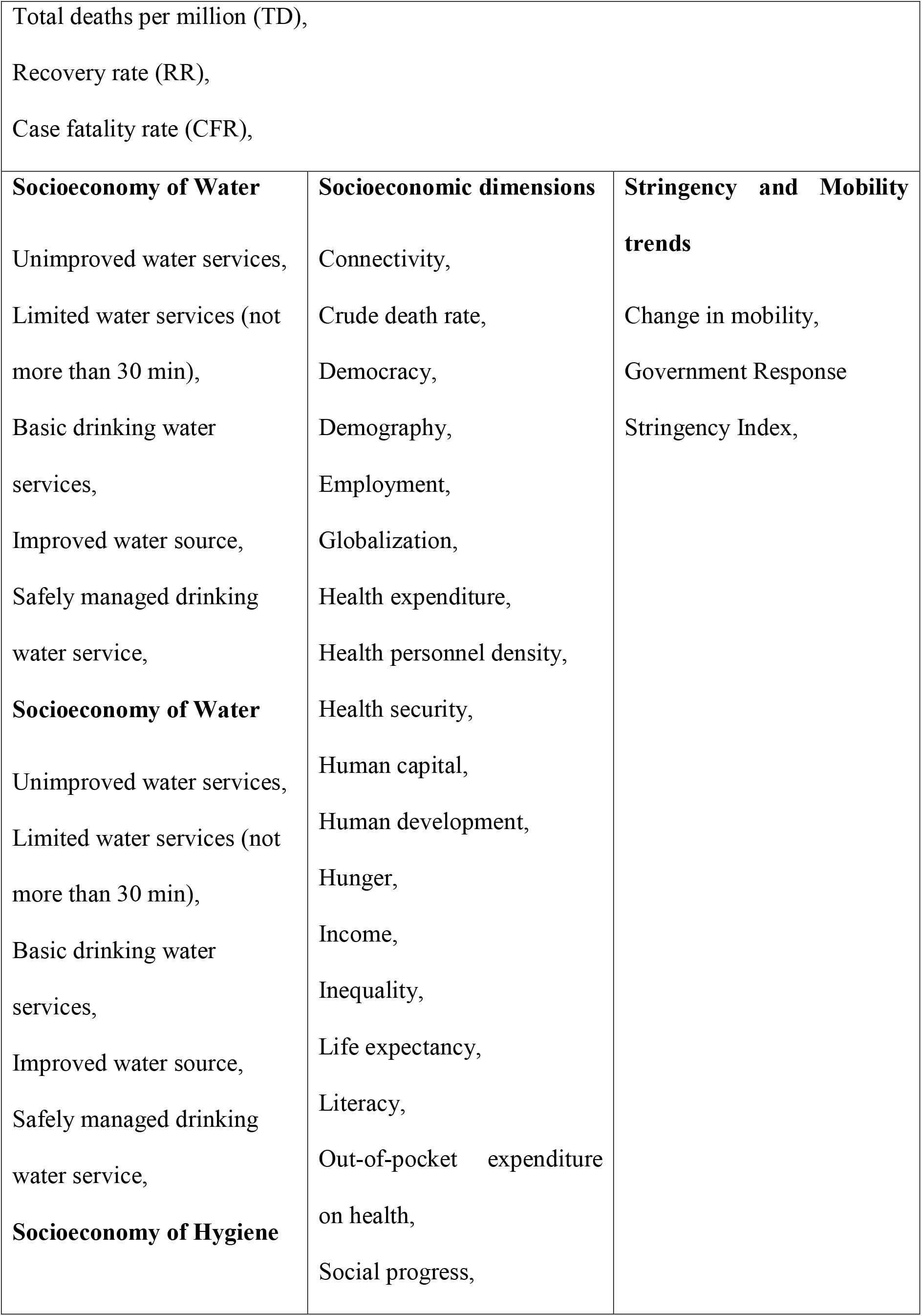
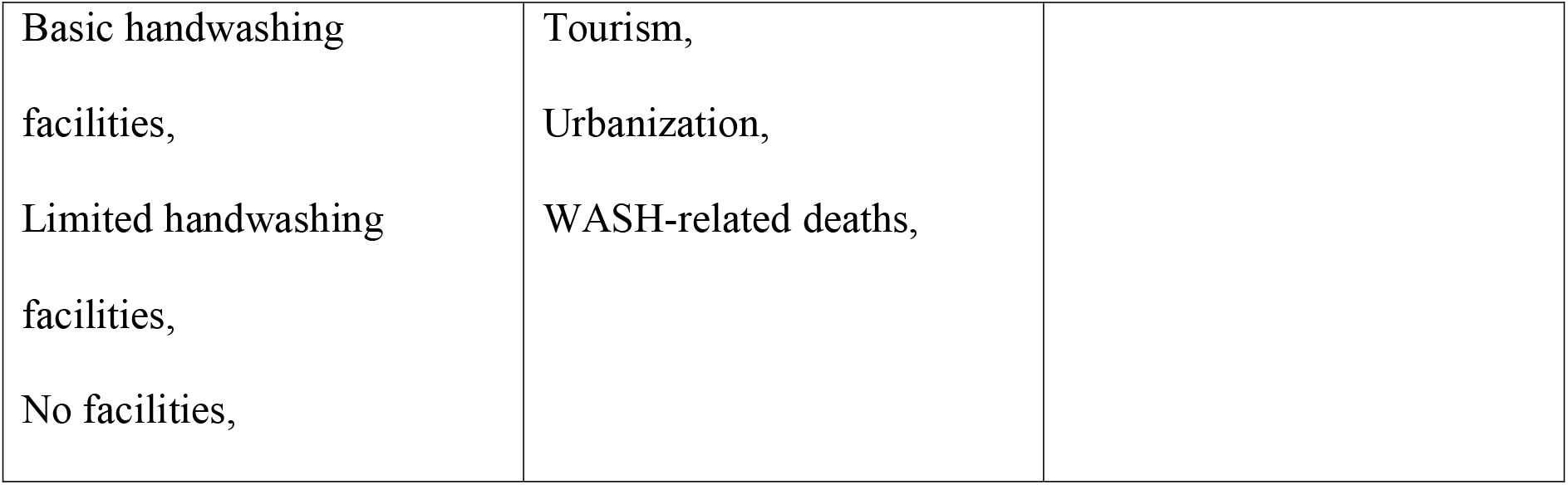
List of indicators used related to various dimensions & Covid-19. (See suppl. file for calculation formula used for RR & CFR)

The statistical analysis has been conducted using R 3.6.2, (R Core Team. 2019) and ‘Hmisc’ (v. 4.4-0) package for performing correlation. Data for worldwide coronavirus cases are taken from package ‘coronavirus’ (v. 0.2.0) from Johns Hopkins University Center for Systems Science and Engineering (JHU CCSE) Coronavirus repository and analysed for cumulative cases for 6 different dates of consecutive 6 months (30-03-2020, 30-04-2020, 30-05-2020, 30-06-2020, 30-07-2020 and 30-08-2020). Though this Covi-19 pandemic started in 31^st^ Dec. 2019, we have taken up March as our starting phase of consideration. Because, the spread of the SARS-CoV2 virus is asymmetrical for the countries of the world, and we can see that from March-April 2020, it had spread to most of the countries of the world with varying degree. We have also selected the span of the duration of study as 6 months (March-August, 2020), because – we think if nothing severely disturbs the dynamics of pandemic (as we can see so far, Nov. 2020), six months is sufficient to convey information about changing influence of various factors (WASH or others) on Covid-19 pandemic. Spearman’s rank-order correlation has been used for finding the association between the variables because the data is not normally distributed. Datasets, used for analysis, is provided in the supplementary files.

## 4. Results

### 4.1. WASH and Covid-19

#### 4.1.1. Water and Covid-19

Most of the indicators that represent good socioeconomic condition related to water, like - basic drinking water, piped services, on-premises services, available when needed services, safely managed drinking water services etc. are strongly positively related to total confirmed, recovered and death. They show a lesser degree of positive correlation with recovery rate and case fatality rate. Another group of indicators that represent not so good socioeconomic condition related to water, like – non-piped services, unimproved water services, limited water services etc. have a strongly negative correlation to total confirmed, recovered and death. They show a lesser degree of negative correlation with recovery rate and case fatality rate.

However, if we delve deeper into each indicator, the see some apparent conflict in results, for all 6 dates (i.e. total span under consideration of the study, 6 months), especially concerning total confirmed (TC) and total death (TD). Some of the indicators that represent good (BDWS) or better (IWS, IWS – P, IWS – AN, IWS - AP) conditions of safe and better access to water are positively related to total confirmed and total death. Also, some of the indicators that represent poor (UWS, LWS) of safe and better access to water are negatively related to total death.

#### 4.1.2. Sanitation and Covid-19

Most of the indicators that represent good socioeconomic condition related to sanitation, like – basic sanitation services, improved sanitation facilities, sewer connection facilities, wastewater treated facilities, septic tank facilities etc. are a higher degree of positive correlation to total confirmed, recovered and death. They show a lesser degree of positive correlation with recovery rate and even lesser with case fatality rate. Another group of indicators that represent not so good socioeconomic condition related to sanitation, like – limited sanitation facilities, open defecation, unimproved sanitation facilities etc. have a strongly negative correlation to total confirmed, recovered and death. They show a less strong negative correlation with recovery rate and even lesser with case fatality rate.

For all 6 dates (i.e. total span under consideration of the study, 6 months), some of the indicators that represent poor (OD, USF, LSF) conditions of safe and better access to sanitation facilities are negatively related to total death and confirmed. Also, indicators that represent good (BSS) or better (ISF, ISF – SC, ISF – ST, ISF – WT, ISF - SM) conditions of safe and better access to water are positively related to total confirmed and total death.

Indicators that depict poor (OD, USF, LSF) conditions of safe and better access to sanitation facilities are negatively related to CFR; whereas indicators that represent good (BSS) or better (ISF, ISF – SC, ISF – WT) conditions of safe and better access to water are positively related to CFR.

#### 4.1.3. Hygiene and Covid-19

Basic handwashing facility has a higher degree of positive correlation to total confirmed, recovered, death and recovery rate. However, it has a lesser degree of positive correlation with the case fatality rate. Another group of hygiene indicator, that represent not so good socioeconomic condition, like – limited and no handwashing facilities have a higher degree of negative correlation with total confirmed, recovered, death and recovery rate. These have a lesser degree of negative correlation with the case fatality rate.

Indicators that represent good (BHF) conditions of safe and better access to hygiene facilities are positively correlated to total confirmed (TC) and total death (TD).

The area marked in red box from the complete correlogram is the interest of this study. Hence, these selected portions, for 6 consecutive months have been displayed in the lower part of this figure.

#### 4.1.4. Trends in WASH-Covid-19 relationships

We can see from Fig 1 that the level of correlation, be it either positive or negative, has decreased over time, i.e. from March to August 2020. This is similar to 3 indicators of Covid-19, namely – total confirmed, recovered and death. Especially, from June 2020 onwards, this is very distinct.

**Fig 1.**
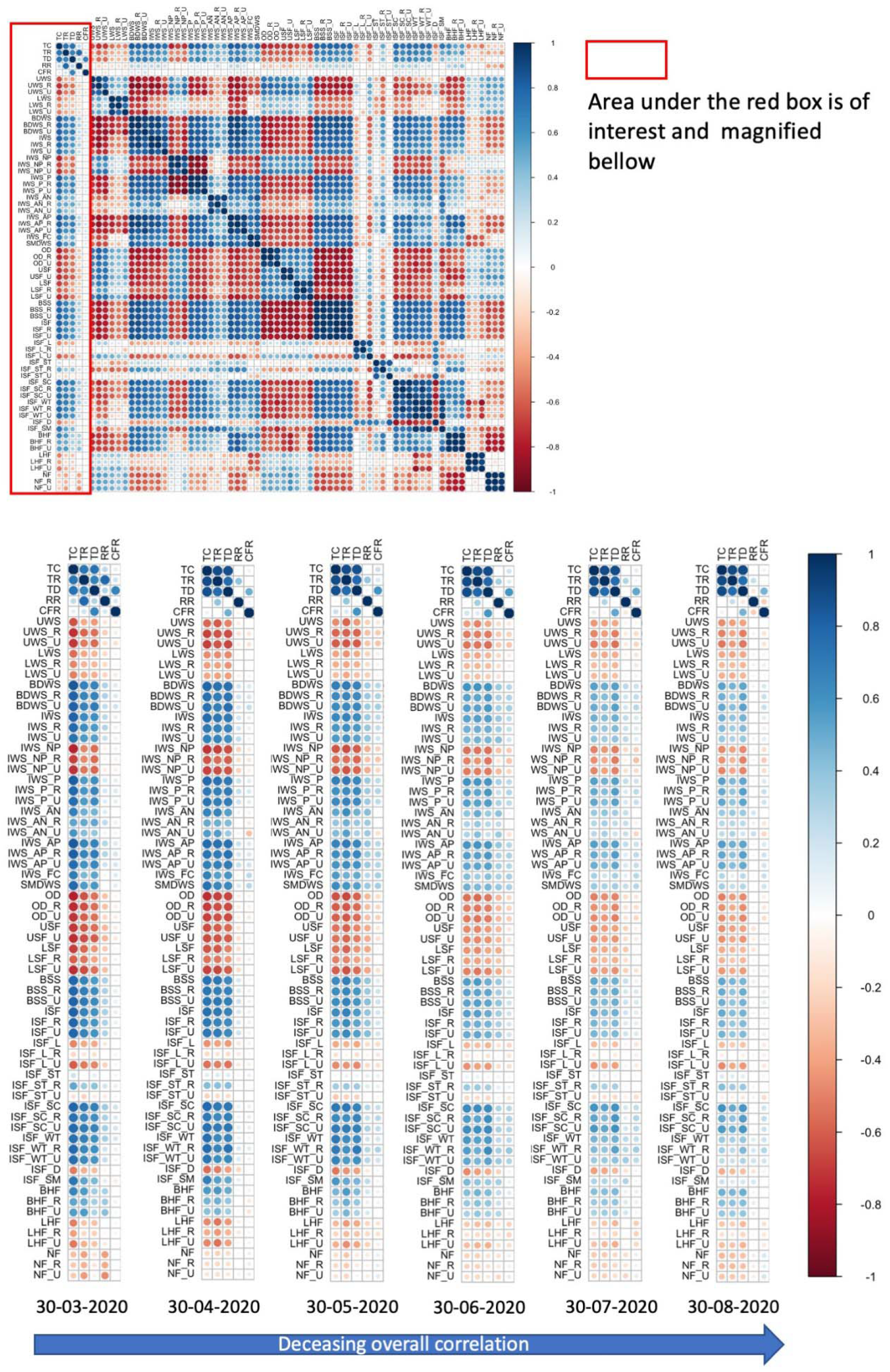
Correlogram from Spearman’s correlation of Covid-19 indicators (n=5) and indicators of water, sanitation and hygiene, WASH access (n=63) for countries (n=175) in the world (March-August, 2020).

From the dendrogram (Suppl. Figure 1) of WASH-Covid-19, it is visible that there have 2 distinct groups throughout the study, one with case fatality and recovery and another with total confirmed, recovered and death. Most of the indicators of WASH have remained polarized in heatmap i.e., indicators of good and bad WASH conditions have remained in two opposite correlation groups in case of one group (with total confirmed, recovered and death). But, for the other group (containing case fatality rate and recovery rate), this is not distinctly different related to WASH indicators.

### 4.2. Socioeconomic dimensions and Covid-19

This study has identified a few contributory elements which have exhibited high association with Covid-19 dynamics across the countries. Among health-related indicators, the share of health expenditure, health security, life expectancy, physician-nurse density and per capita out-of-pocket health expenditure have a higher degree of positive correlation with total confirmed, recovered and death related to Covid-19. Two other health-related indicators have a lesser degree of positive correlation with the same, namely – per capita share of health expenditure and crude death rate. Share of per capita out-of-pocket health expenditure has a higher degree of negative correlation with the same 3 indicators related to Covid-19.

Among economy-related indicators, globalization, real GDP and connectedness have a higher degree of positive correlation with total confirmed, recovered and death related to Covid-19. But the Gini coefficient has a lesser degree of negative correlation with the same.

Among demographic indicators, the median age of the population and aged population have a higher degree of positive correlation than population density with total confirmed, recovered and death; whereas the total population is negatively correlated with them.

Among societal indicators, literacy rate, democracy level, human capital and development, international tourism arrival, the share of urban population and social progress have a higher degree of positive correlation than with total confirmed, recovered and death; whereas employment to population ratio, hunger level, the share of vulnerable employment, unemployment and prevalence of deaths from water, sanitation and handwashing facilities have a negative correlation with the same.

The area marked in red box from the complete correlogram is the interest of this study. Hence, these selected portions, for 6 consecutive months have been displayed in the lower part of this figure.

#### 4.2.1. Trends in Socioeconomy-Covid-19 relationship

From the dendrogram (Suppl. Figure 2) of socioeconomy-Covid-19, it is clearly visible that there have 2 distinct groups throughout the study, one with case fatality and recovery and another with the rest (i.e. total confirmed, recovered and death).

**Fig 2.**
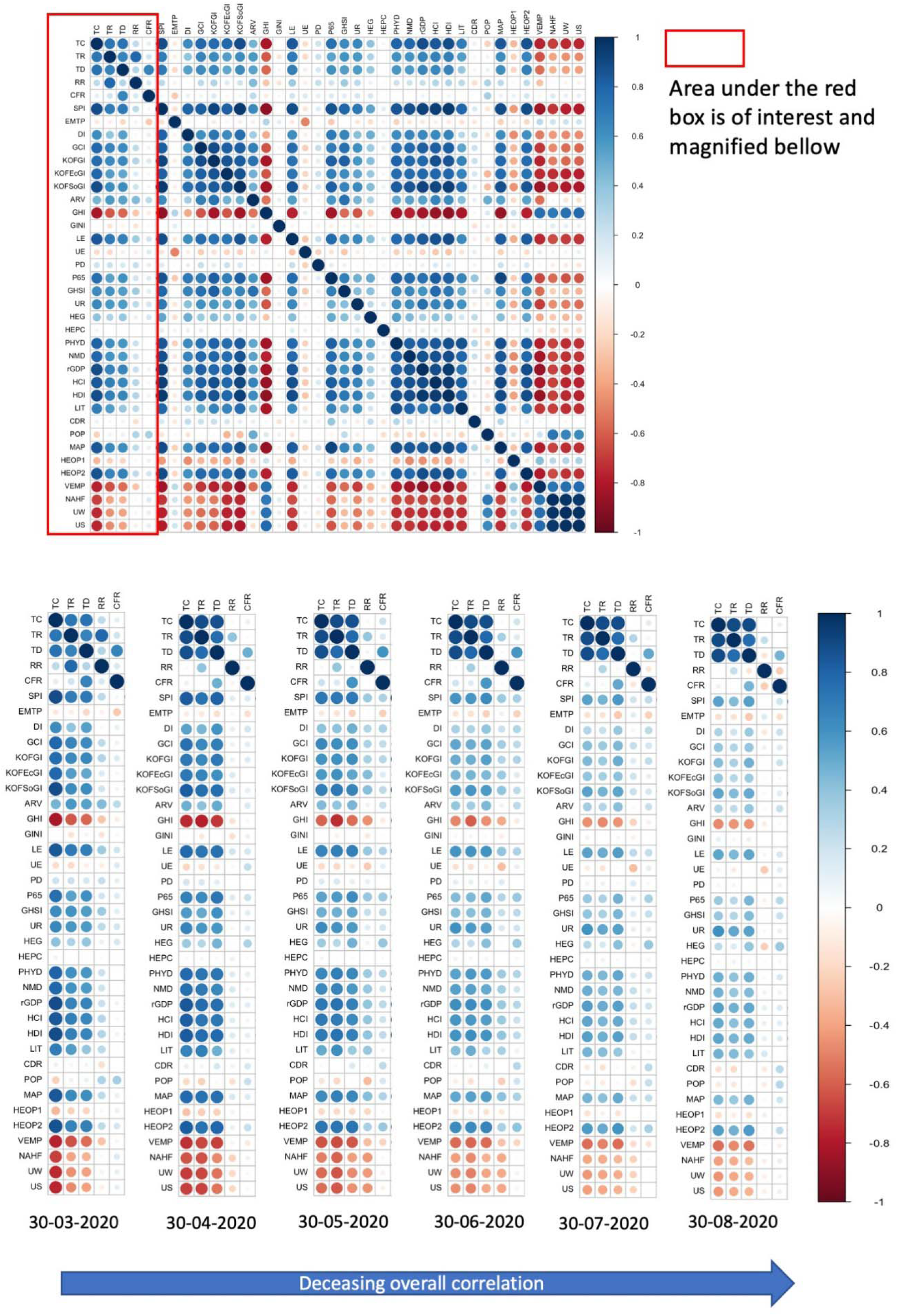
Correlogram from Spearman’s correlation of Covid-19 indicators (n=5) and indicators of socio-economic situations (n=33) for countries (n=175) in the world (March-August, 2020).

Mostly, the indicators of socioeconomic conditions have remained polarized in heatmap i.e., indicators of improved and poor socioeconomic conditions have remained in two opposite correlation groups in case of one group (with total confirmed, recovered and death). But, for the other group (containing case fatality rate and recovery rate), this is not distinctly different related to socioeconomic indicators.

### 4.3. Stringency, mobility and Covid-19

Among these indicators, time spent in residential areas (i.e. Rsd) has a higher degree of positive correlation with total confirmed, recovered and death related to Covid-19, than stringency index (SnI). On the other hand, the density of visitors in workplaces (WkP), transit stations (TnS) and retail & recreation places (RnR) have a higher degree of negative correlation to those indicators of Covid-19, than the density of visitors in grocery, pharmacy (GnP) and parks (Pks). Primarily (i.e. March 2020), the case fatality rate is more correlated to these indicators that recovery rate, and both have a lesser degree of correlation that aforementioned 3 indicators regarding Covid-19. Excluding the first month (March 2020), the recovery rate has been negatively correlated to stringency.

#### 4.3.1. Trends in Stringency and mobility - Covid-19 relationships

From the dendrogram (Suppl. Figure 3) of stringency-mobility-Covid-19, it is clearly visible that there have 2 distinct groups during the earlier stage of study (March-May 2020), one with case fatality and another with the rest (i.e. recovery, total confirmed, recovered and death). However, its nature has changed in the following months (June – August 2020), one group with case fatality and recovery and another group with the rest (i.e. total confirmed, recovered and death).

**Fig 3.**
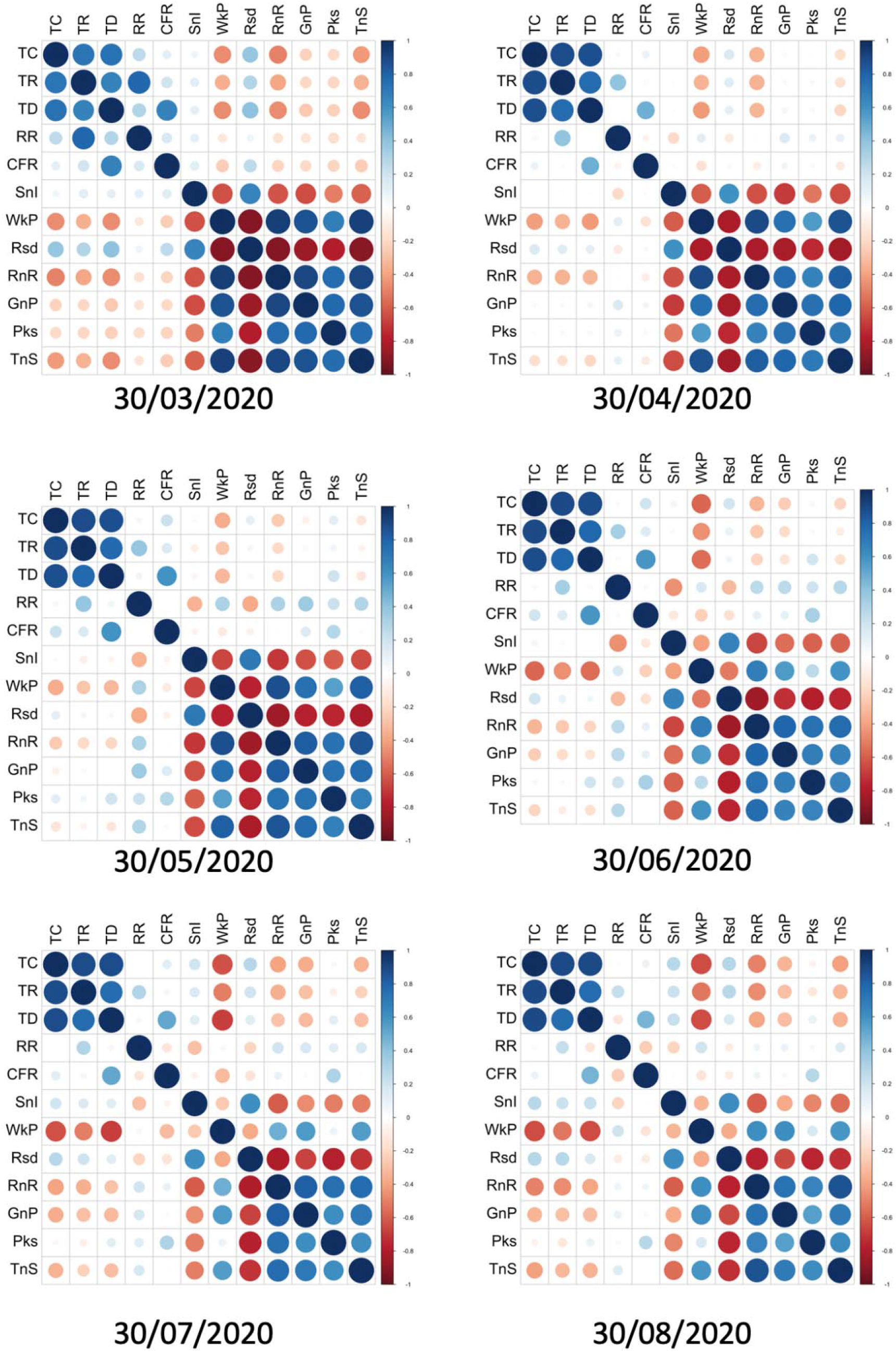
Correlogram from Spearman’s correlation of Covid-19 indicators (n=5) and indicators of socio-economic situations (n=7) for countries (n=175) in the world (March-August, 2020).

Mostly, the indicators of stringency and mobility conditions have not remained polarized in heatmap i.e., the nature and degree of correlation are changing with time.

The overall time wise (here, month) change of number of significantly correlated associations between Covid-19 and other 3 groups of indicators (viz. WASH, socioeconomic factors and stringency & mobility) are shown in Fig 4.

**Fig 4.**
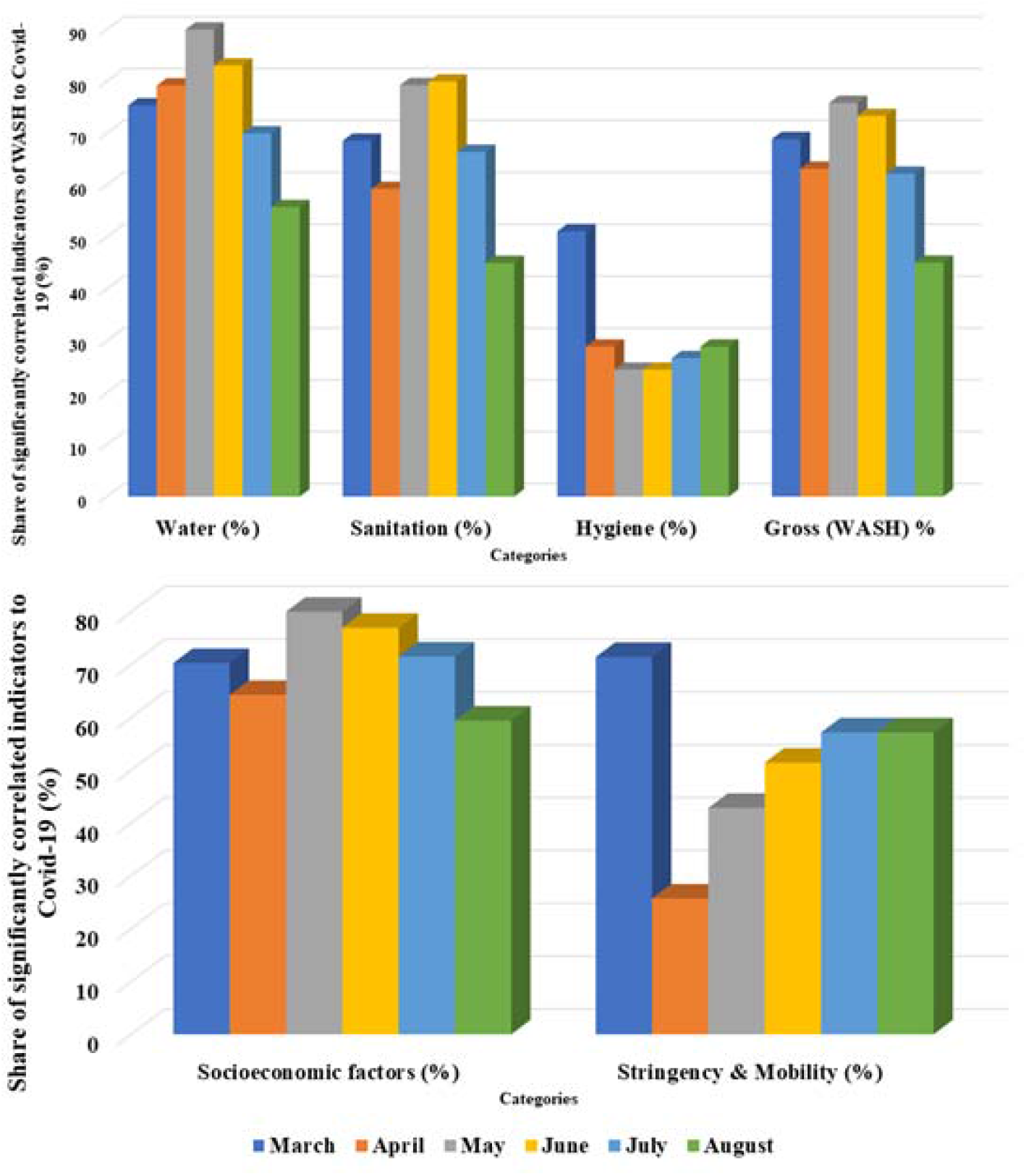
Temporal change (month wise) in the number of significantly correlated indicators in association with Covid-19 pandemic. (See Suppl. Table 6 for underlying data from calculation)

## 5. Discussion

From this study, there are some basic understanding that can be inferred. For the WASH sphere, providing basic as well as improved drinking water, sanitation and hygiene facilities are directly positively connected to the recovery rate of Covid-19. Likewise, reducing unimproved drinking water, sanitation and open defecation is negatively connected to recovery rate. This means, not only for the specific domain of WASH but inclusive access to these facilities are also connected to the health sector as well as disaster prevention (like – a pandemic connected to water). The hassle-free access of purified water (at least not associated to wastewater, Lapolla et al., 2020; Bogler et al, 2020) at home premises (Frumkin et al., 2020; Kassem and Jaafar, 2020; Staddon et al., 2020), access to improved sanitation facilities at household level as well as the availability of household hygiene measures (in form of soaps, hand sanitizers etc.) (Schmidt, 2020; Yu et al., 2020) have become the frontier force of precautionary prevention.

The other socio-economic factors also ascribe to similar perception regarding the covid-19 pandemic. Among the health-related factors, better health security, a higher density of physicians and nurses, better life expectancy, higher per capita out of pocket expenditure and the lower crude death rate is positively associated with better recovery rate. These results are in line with Adams & Walls, 2020 (for healthcare workforce), Kandel et al., 2020 (for health security) etc. Among the economic factors, higher real GDP, connectedness, globalization and lower economic inequality are positively associated with a better rate of recovery. These outcomes are in line with Makridis & Hartley, 2020 (for real GDP), McKibbin & Fernando, 2020 (for macroeconomy) etc. Among the societal factors, higher literacy, social progress, democracy score, human capital and development along with lower unemployment, employment vulnerability and huger are associated with better recovery. These findings are also in line with UNDP report (for human development), Khemraj & Yu, 2020 (for human capital), Kawohl & Nordt, 2020 (for unemployment) etc (see Fig 5).

**Fig 5.**
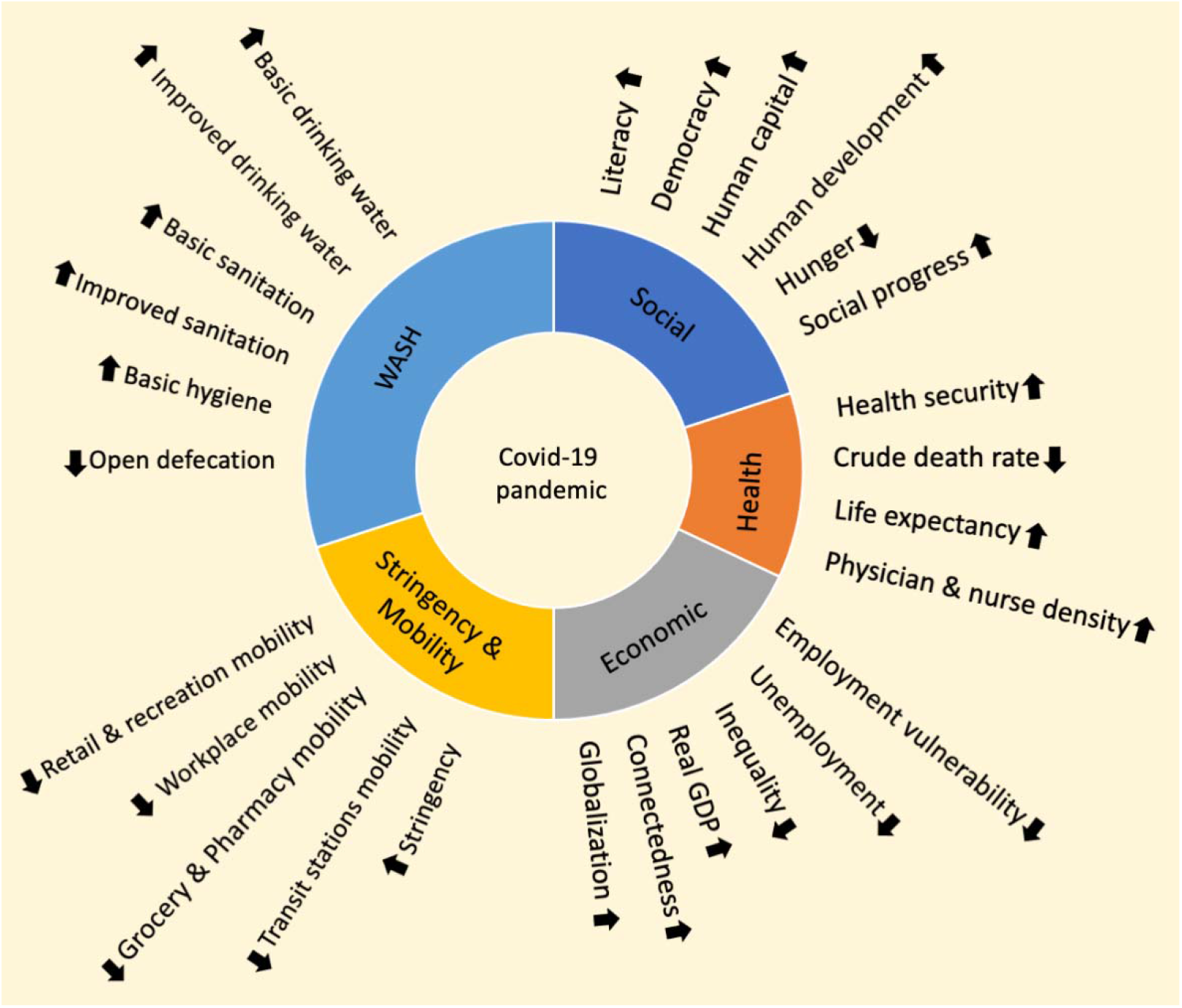
An overall global pandemic preparedness as well as mitigation actions that need to be given significance as the outcome established from this study.

This new pandemic has undoubtedly emphasised the frivolity of the present condition as well as the significance of the global WASH access system. These are the bare minimum requirements which have been either neglected or not properly implemented (sometimes due to lack of awareness). Other socioeconomic factors, though of varying degree, has a high level of correlation with various indicators of the covid-19 pandemic. This entails towards the importance of deliverance of socio-economic development globally.

There are some possible drawbacks in our work. Firstly, WASH data is from 2017, not immediate pre-pandemic data. It is because WHO-UNICEF JMP dataset is most robust and uniform till date. However, recently, there have been some issues regarding the applicability of WASH data towards predicting the WASH-Covid19 relationship accurately (Quispe-Coica and Pérez-Foguet, 2020). But we don’t have any better alternative. Second, the socio-economic indicators are also not of immediately pre-pandemic time (see Suppl. Table 5). This is due to the unavailability of the latest data (2019-20) for all of the socioeconomic indicators. Third, this analysis has not been done on the sub-national or lower scale. Because the objective was to understand the determinants of the covid-19 pandemic in general, for large scale. Also, the sub-national data are not comprehensive and uniformly available for all countries. Fourth, the study period (March-August 2020, i.e. 6 months) is odd. Though the onset and spread of covid-19 pandemic are asymmetric for different countries, we think from March 2020 onwards, it had started in most of the major countries of the world. Also, we have taken 6 months (up to August 2020) because the study also tries to understand and lingering association of determinants with Covid-19 pandemic, and as we can see from the data that this duration is long enough to serve the purpose. Lastly, we have chosen only one date (30^th^ of each month) which is not continuous. We have taken nearly equidistant dates from each month as we think this is enough to give us a broad outline of the role of determinants.

We see whatever kind of association do the determining factors have with those of Covid-19, the degree is lessening through time (except – those related to mobility and stringency). This means two things, (a) other factors (which are outside the operational boundary of this study) might be getting superior control over the dynamics of Covid-19 than those included in this study, and/or (b) the data of aforementioned determinants are of pre-pandemic time, means they are not being updated daily or monthly (like – Covid-19). From this we can infer, these factors have been determining the trajectory of the covid-19 pandemic in the old normal (pre-pandemic) as well as will continue to do so in the new normal (post-pandemic). We think once robust data are available of these determinants after the pandemic, we might see a similar level of association.

The present study has advocated the utility of interpreting determinants in epidemiological research, which can be diverse, with this Covid-19 pandemic as a case of example. The present work has explored the association between the WASH, social, demographic, mobility, economic as well as health parameters and Covid-19, extracting insightful tales of dynamics of Covid-19 pandemic. Therefore, we hope, this study could be a reference for future health research and their controlling factors towards synthesizing a multidisciplinary approach for either prevention or control or both (Ross et al., 2020). Additionally, all the data used in this work are open source and freely available (we have also provided the raw data and results in supplement), in so doing, reproducibility, as well as scientific replicability, can be achievable.

## Data Availability

Data will be made available upon request from Corresponding author.

