## Supplementary file for "Water, Sanitation, Hygiene (WASH) or others? - a global analysis on determinants of Covid-19 pandemic"

**Heatmaps:**

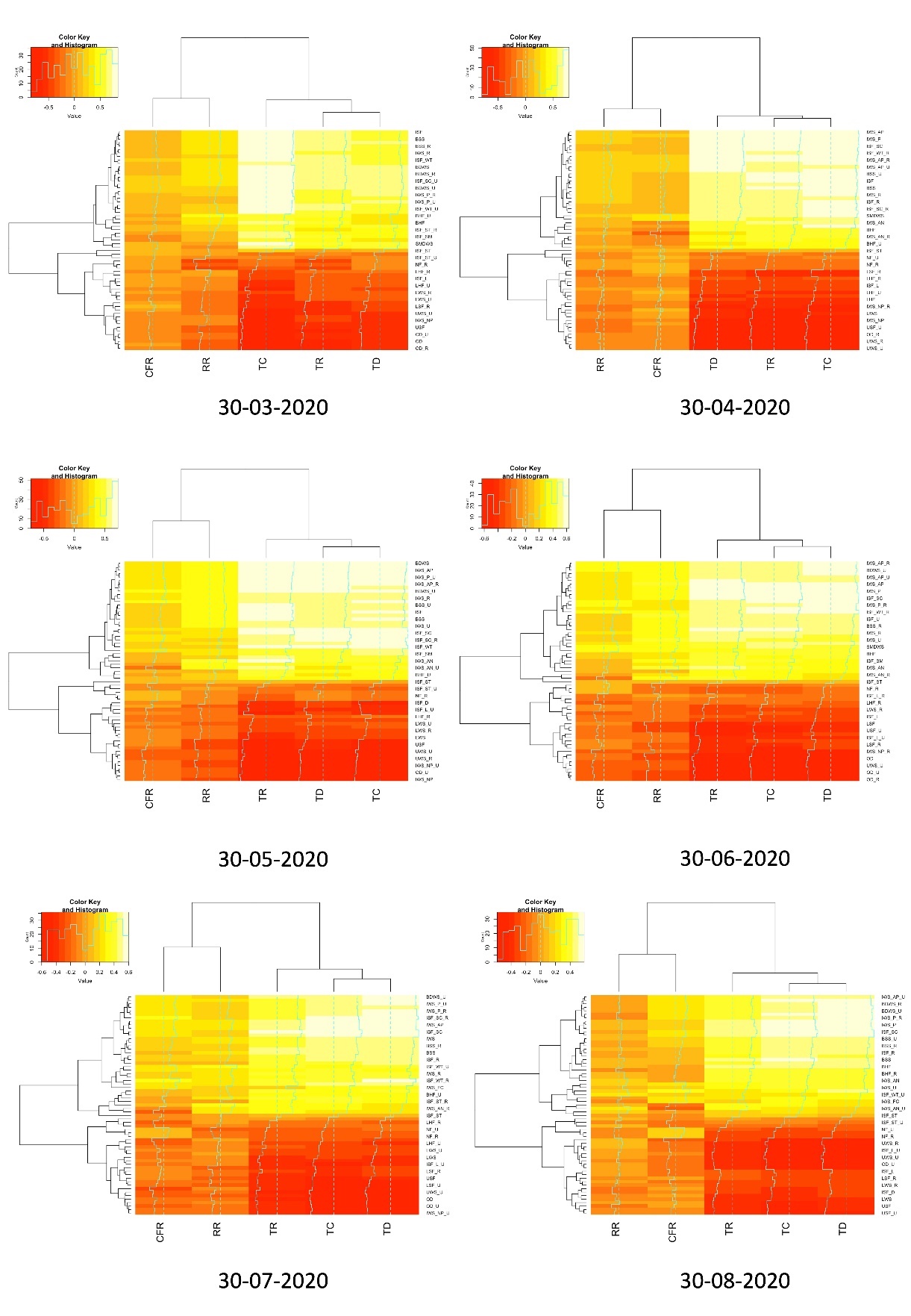

Fig 1. Heatmap of correlation between various indicators of WASH and Covid-19 for 6 months (March-August 2020).

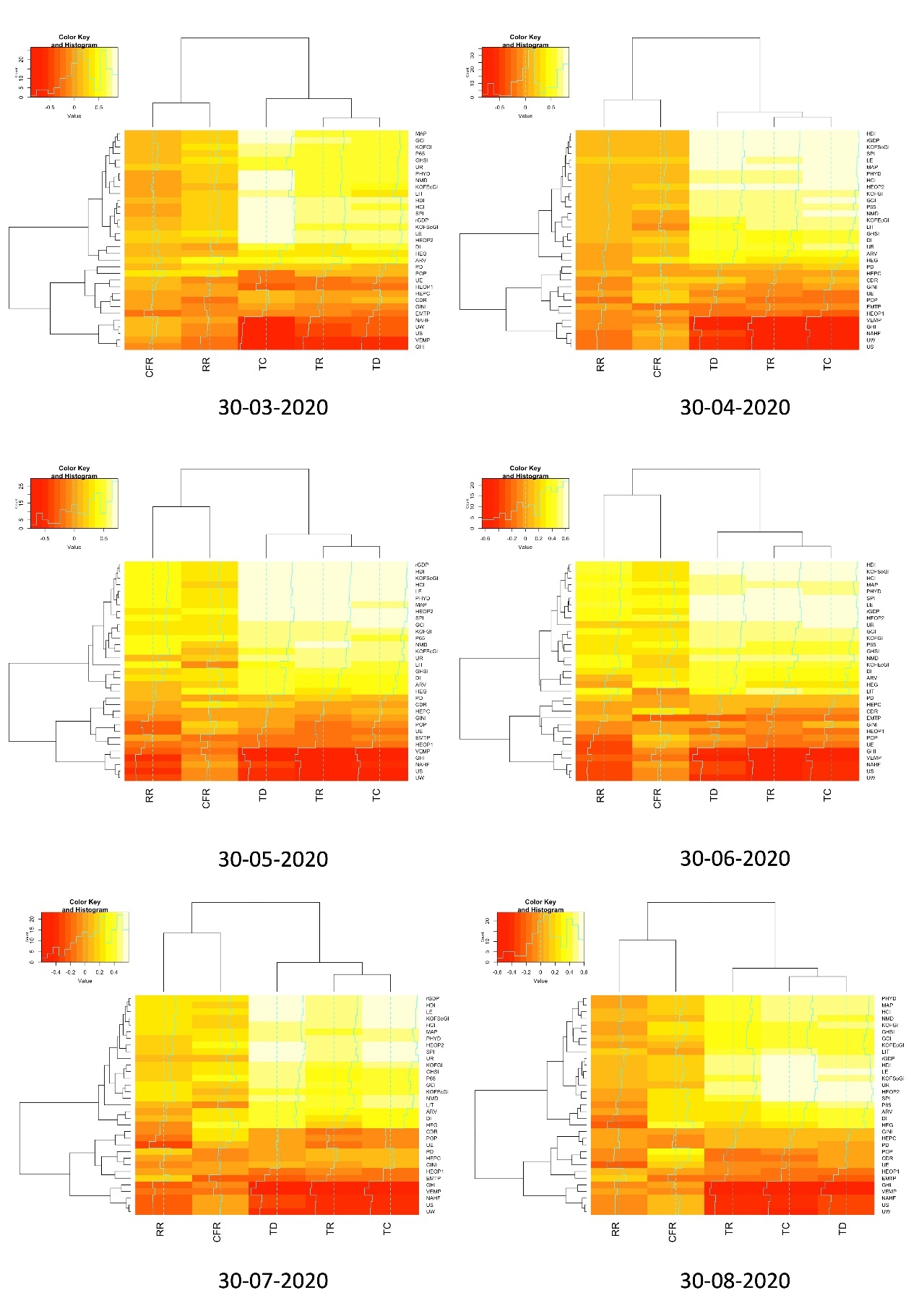

Fig 2. Heatmap of correlation between various indicators of socio-economy (excluding WASH) and Covid-19 for 6 months (March-August 2020).

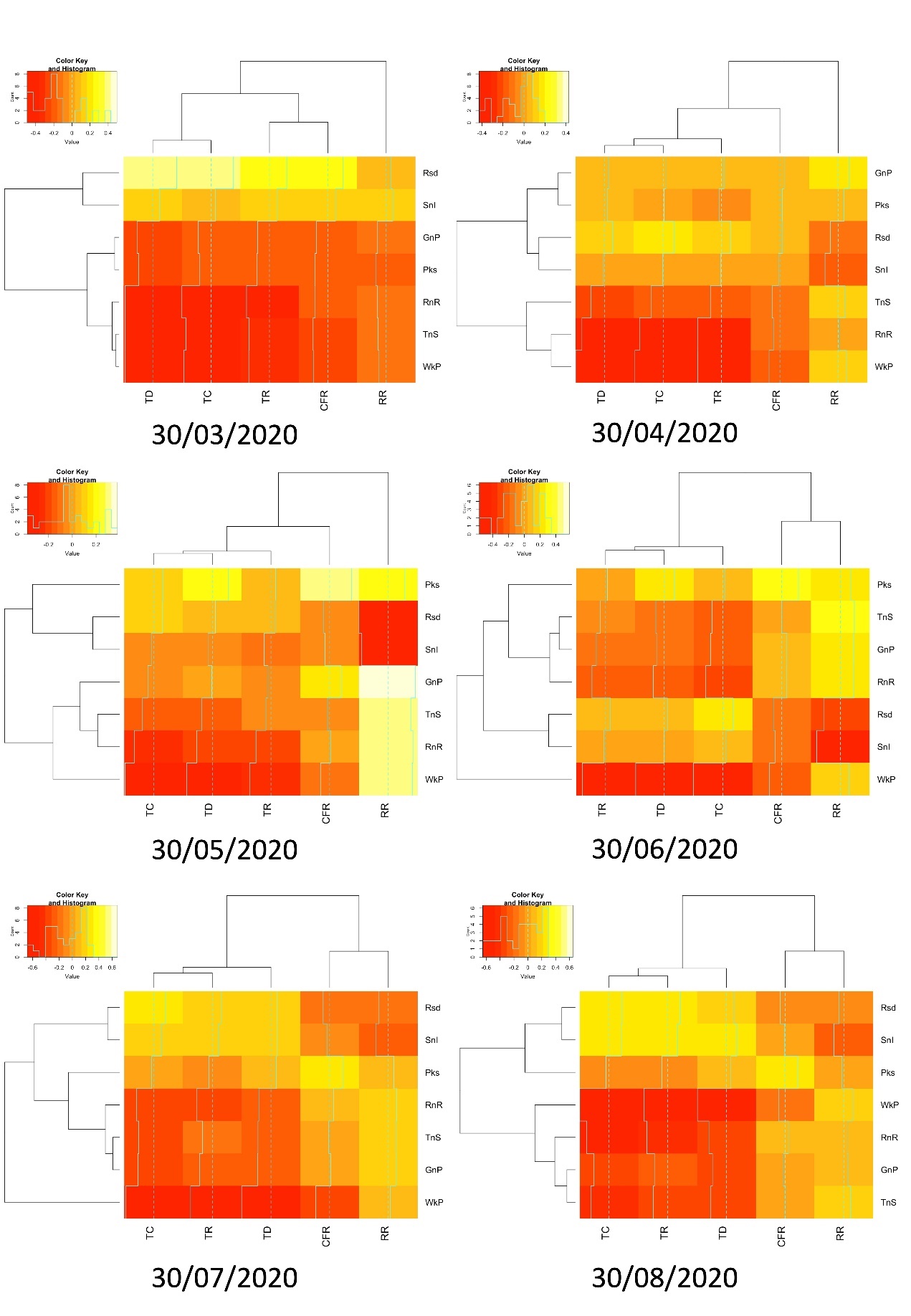

Fig 3. Heatmap of correlation between various indicators of stringency & mobility and Covid-19 for 6 months (March-August 2020).

Calculation of indicators of Covid-19:

Recovery rate (RR) = (total recovered/total confirmed) ×100

Case fatality rate (CFR) = (total death/ total confirmed) ×100

Connection of Water to UN – SDG:

Target 6.1, 'By 2030, achieve universal and equitable access to safe and affordable drinking water for all'.

Table 1. Indicators for socioeconomy of Water

| **No.** | **Indicators** | **Abbreviation** | **No.** | **Sub-indicators** |
| --- | --- | --- | --- | --- |
| 1 | Unimproved water services | UWS | 1 | overall (% of the population) |
|  |  | UWS - R | 2 | rural (% of rural population) |
|  |  | UWS - U | 3 | urban (% of urban population) |
| 2 | Limited water services (not more than 30 min) | LWS | 4 | overall (% of the population) |
|  |  | LWS - R | 5 | rural (% of rural population) |
|  |  | LWS - U | 6 | urban (% of urban population) |
| 3 | Basic drinking water service | BDWS | 7 | overall (% of the population) |
|  |  | BDWS - R | 8 | rural (% of rural population) |
|  |  | BDWS - U | 9 | urban (% of urban population) |
| 4 | Improved water source | IWS | 10 | overall (% of the population with access) |
|  |  | IWS – R | 11 | rural (% of rural population with access) |
|  |  | IWS – U | 12 | urban (% of urban population with access) |
| 5 | Improved water services, non-piped | IWS - NP | 13 | overall (% of population) |
|  |  | IWS - NP - R | 14 | rural (% of rural population) |
|  |  | IWS - NP - U | 15 | urban (% of urban population) |
| 6 | Improved water services, piped | IWS - P | 16 | overall (% of the population) |
|  |  | IWS - P - R | 17 | rural (% of rural population) |
|  |  | IWS - P - U | 18 | urban (% of urban population) |
| 7 | Improved water services, available when needed | IWS - AN | 19 | overall (% of the population) |
|  |  | IWS - AN - R | 20 | rural (% of rural population) |
|  |  | IWS - AN - U | 21 | urban (% of urban population) |
| 8 | Improved water services, accessible on-premises | IWS - AP | 22 | overall (% of the population) |
|  |  | IWS - AP - R | 23 | rural (% of rural population) |
|  |  | IWS - AP - U | 24 | urban (% of urban population) |
| 9 | Improved water services, free from contamination | IWS - FC | 25 | overall (% of population) |
| 10 | Safely managed drinking water service | SMDWS | 26 | overall (% of the population) |

Connection of Sanitation to UN – SDG:

Target 6.2 "by 2030, achieve access to adequate and equitable sanitation and hygiene for all and end open defecation, paying special attention to the needs of women and girls and those in vulnerable situations"

Table 2. Indicators for socioeconomy of Sanitation

| **No.** | **Indicators** | **Abbreviation** | **No.** | **Sub-indicators** |
| --- | --- | --- | --- | --- |
| 1 | Open defecation | OD | 1 | overall (% of the population) |
|  |  | OD - R | 2 | rural (% of rural population) |
|  |  | OD - U | 3 | urban (% of urban population) |
| 2 | Unimproved sanitation facilities | USF | 4 | overall (% of the population) |
|  |  | USF - U | 5 | urban (% of urban population) |
| 3 | Limited (shared) sanitation facilities | LSF | 6 | overall (% of the population) |
|  |  | LSF - R | 7 | rural (% of rural population) |
|  |  | LSF - U | 8 | urban (% of urban population) |
| 4 | Basic sanitation service | BSS | 9 | overall (% of population) |
|  |  | BSS - R | 10 | rural (% of rural population) |
|  |  | BSS - U | 11 | urban (% of urban population) |
| 5 | Improved sanitation facilities | ISF | 12 | overall (% of the population) |
|  |  | ISF - R | 13 | rural (% of rural population) |
|  |  | ISF - U | 14 | urban (% of urban population) |
| 6 | Improved sanitation facilities (excluding shared) - Latrines and other | ISF - L | 15 | overall (% of the population) |
|  |  | ISF - L - R | 16 | rural (% of rural population) |
|  |  | ISF - L - U | 17 | urban (% of urban population) |
| 7 | Improved sanitation facilities (excluding shared) - Septic tanks | ISF - ST | 18 | overall (% of the population) |
|  |  | ISF - ST - R | 19 | rural (% of rural population) |
|  |  | ISF - ST - U | 20 | urban (% of urban population) |
| 8 | Improved sanitation facilities (excluding shared) - Sewer connections | ISF - SC | 21 | overall (% of population) |
|  |  | ISF - SC - R | 22 | rural (% of rural population) |
|  |  | ISF - SC - U | 23 | urban (% of urban population) |
| 9 | Improved sanitation facilities (excluding shared) - Wastewater treated | ISF - WT | 24 | overall (% of the population) |
|  |  | ISF - WT - R | 25 | rural (% of rural population) |
|  |  | ISF - WT - U | 26 | urban (% of urban population) |
| 10 | Improved sanitation facilities - Disposed in situ | ISF - D | 27 | overall (% of population) |
| 11 | Improved sanitation facilities - Safely managed | ISF - SM | 28 | overall (% of population) |

Table 3. Indicators for socioeconomy of Hygiene

| **No.** | **Indicators** | **Abbreviation** | **No.** | **Sub-indicators** |
| --- | --- | --- | --- | --- |
| 1 | Basic handwashing facilities | BHF | 1 | overall (% of the population) |
|  |  | BHF - R | 2 | rural (% of rural population) |
|  |  | BHF - U | 3 | urban (% of urban population) |
| 2 | Limited (without water or soap) handwashing facilities | LHF | 4 | overall (% of the population) |
|  |  | LHF - R | 5 | rural (% of rural population) |
|  |  | LHF - U | 6 | urban (% of urban population) |
| 3 | No facilities | NF | 7 | overall (% of the population) |
|  |  | NF - R | 8 | rural (% of rural population) |
|  |  | NF - U | 9 | urban (% of urban population) |

Table 4. Indicators of socioeconomic dimensions

| **No.** | **Indicator** | **Type** | **Abbreviation** |
| --- | --- | --- | --- |
| 1 | Adult Literacy rate (% of people ages ≥15y) (2016-18) * | Society | LIT |
| 2 | Current health expenditure (% of GDP) (2017) | Health | HEG |
| 3 | Current health expenditure per capita, current US$ (2017) | Health | HEPC |
| 4 | Death rate, crude (per 1,000 people) (2018) | Health | CDR |
| 5 | Democracy index (2019) | Society | DI |
| 6 | Economic globalization Index (2017) | Economy | KOFEcGI |
| 7 | Employment to population ratio, 15y+ (%) (2019) | Society | EMTP |
| 8 | GDP per capita (constant 2010 US$) (2017-19) * | Economy | rGDP |
| 9 | Gini coefficient (2020) | Economy | GINI |
| 10 | Global connectedness index (2017) | Economy | GCI |
| 11 | Global health security index (2019) | Health | GHSI |
| 12 | Global hunger index (2019) | Society | GHI |
| 13 | Globalization index (2017) | Economy | KOFGI |
| 14 | Human capital index (2020) | Society | HCI |
| 15 | Human development index (2018) | Society | HDI |
| 16 | International tourism, number of arrivals (2017-18) * | Society | ARV |
| 17 | Life expectancy (2020) | Health | LE |
| 18 | The median age of the population, years (2020) | Demography | MAP |
| 19 | No access to handwashing facility (number of deaths) (2017) | Society | NAHF |
| 20 | Nurses and midwives (per 1,000 people) (2016-18) * | Health | NMD |
| 21 | Out-of-pocket expenditure (% of current health expenditure) (2017) | Health | HEOP1 |
| 22 | Out-of-pocket expenditure per capita (current US$) (2017) | Health | HEOP2 |
| 23 | Physicians (per 1,000 people) (2016-18) * | Health | PHYD |
| 24 | Population density (2019) | Demography | PD |
| 25 | Population ages ≥ 65 years (% of total) (2019) | Demography | P65 |
| 26 | Population, number (2020) | Demography | POP |
| 27 | Share of vulnerable employment (% of total) (2019) | Society | VEMP |
| 28 | Share of urban population (% of total) (2019) | Society | UR |
| 29 | Social globalization Index (2017) | Economy | KOFSoGI |
| 30 | Social progress index (2019) | Society | SPI |
| 31 | Unemployment rate (2020) | Society | UE |
| 32 | Unsafe sanitation (number of deaths) (2017) | Society | US |
| 33 | Unsafe water source (number of deaths) (2017) | Society | UW |

* Using latest year of data available.

Table 5. Google Community Mobility Reports (CMR) definitions

| **No.** | **Indicator** | **Abbreviation** | **Description** |
| --- | --- | --- | --- |
| 1 | Grocery & Pharmacy | GnP | Mobility trends for places like grocery markets, food warehouses, farmers markets, speciality food shops, drug stores, and pharmacies. |
| 2 | Parks | Pks | Mobility trends for places like local parks, national parks, public beaches, marinas, dog parks, plazas, and public gardens. |
| 3 | Residential | Rsd | Mobility trends for places of residence. |
| 4 | Retail & recreation | RnR | Mobility trends for places like restaurants, cafes, shopping centres, theme parks, museums, libraries, and movie theatres. |
| 5 | Transit stations | TnS | Mobility trends for places like public transport hubs such as subway, bus, and train stations. |
| 6 | Workplaces | WkP | Mobility trends for places of work. |

Table 6. Change in the number of significantly correlated indicators

| **No.** | **Date** | **Water (%)** | **Sanitation (%)** | **Hygiene (%)** | **Gross (%)** | **Socioeconomic factors (%)** | **Stringency & Mobility (%)** |
| --- | --- | --- | --- | --- | --- | --- | --- |
| 1 | 30-03-2020 | 75.38 | 68.57 | 51.11 | 68.89 | 70.3 | 71.43 |
| 2 | 30-04-2020 | 79.23 | 59.28 | 28.89 | 63.17 | 64.24 | 25.71 |
| 3 | 30-05-2020 | 90 | 79.28 | 24.44 | 75.87 | 80 | 42.86 |
| 4 | 30-06-2020 | 83.08 | 80 | 24.44 | 73.33 | 76.97 | 51.43 |
| 5 | 30-07-2020 | 70 | 66.43 | 26.66 | 62.22 | 71.51 | 57.14 |
| 6 | 30-08-2020 | 50.77 | 45 | 28.88 | 45.07 | 59.39 | 57.14 |

**Data Sources**

- Altman, S. A., Ghemawat, P., Bastian, P. 2019. DHL Global Connectedness Index 2018. Deutsche Post DHL. (<https://www.dhl.com/global-en/home/insights-and-innovation/thought-leadership/case-studies/global-connectedness-index.html>)
- Democracy index (<https://www.eiu.com/topic/democracy-index>)
- Global health security index (<https://www.ghsindex.org/>)
- Global hunger index (<https://www.globalhungerindex.org/results.html>)
- Gygli, S., Haelg, F., Potrafke, N., Sturm, J-E. (2019). The KOF Globalisation Index – revisited. The Review of International Organizations, 14, 543–574. <https://doi.org/10.1007/s11558-019-09344-2>
- Human capital index (<https://www.worldbank.org/en/publication/human-capital>)
- Human development index (<http://hdr.undp.org/en/data>)
- Our World in Data (<https://ourworldindata.org/policy-responses-covid>)
- Social Progress Index (<https://www.socialprogress.org/>)
- WHO/UNICEF JMP (WHO/UNICEF Joint Monitoring Programme for Water Supply, Sanitation and Hygiene). 2020. (<https://washdata.org/>)
- World Bank, 2020. World Development Indicators. <http://data.worldbank.org/>
- World Population Review (<https://worldpopulationreview.com/>)
